# Projecting the impact of nutrition policy to improve child stunting: A case study in Guatemala using the Lives Saved Tool

**DOI:** 10.1101/2021.05.24.21256641

**Authors:** Scott Tschida, Ana Cordon, Gabriela Asturias, Mónica Mazariegos, María F Kroker-Lobos, Bianca Jackson, Peter Rohloff, David Flood

## Abstract

**Background:** Child stunting is a critical global health issue. Guatemala has one of the world’s highest levels of stunting despite sustained commitment to international nutrition policy best-practices endorsed by the Scaling Up Nutrition movement (SUN). Our objective was to use Guatemala as a case study by projecting the impact of a recently published national nutrition policy, the Great Crusade, that is consistent with SUN principles.

**Methods:** We used the Lives Saved Tool (LiST) to project the scaling-up of nutrition interventions proposed in the Great Crusade and recommended by SUN. Our outcomes were changes in stunting prevalence, number of stunting cases averted, and number of cases averted by intervention in children under five years of age from 2020-2030. We considered four scenarios: (1) intervention coverage continues based on historical trends, (2) coverage targets in the Great Crusade are achieved, (3) coverage targets in the Great Crusade are achieved with reduced fertility risk, and (4) coverage reaches an aspirational level.

**Results:** All scenarios led to modest reductions in stunting prevalence. In 2024, stunting prevalence was estimated to change by -0.1% (95% CI 0.0% to -0.2%) if historical trends continue, -1.1% (95% CI -0.8% to -1.5%) in the Great Crusade scenario, and -2.2% (95% CI - 1.6% to -3.0%) in the aspirational scenario. In 2030, we projected a stunting prevalence of -0.4% (95% CI -0.2% to -0.8%) and -3.7% (95% CI -2.8% to -5.1%) in the historical trends and aspirational scenario, respectively. Complementary feeding, sanitation, and breastfeeding were the most impactful interventions across models.

**Conclusions:** Targeted reductions in child stunting prevalence in Guatemala are unlikely to be achieved solely based on increases in intervention coverage. Our results show the limitations of current paradigms recommended by the international nutrition community. Policies and strategies are needed that address the broader structural drivers of stunting.

## BACKGROUND

More than 149 million children under five years of age worldwide have inadequate linear growth, which is also referred to as child stunting.^1^ Stunting is a crucial global child health issue given its negative impacts on health and well-being throughout the lifespan. In the short-term, stunted children are more likely to experience morbidity, mortality, and developmental delays.^2^ In the long-term, adults who were stunted as children are at risk for lower cognitive performance, educational achievement, and economic productivity.^3–5^ Stunting also may contribute to the development of adult cardiometabolic conditions such as diabetes, obesity, and hypertension.^6,7^ Sustainable Development Goal (SDG) target 2.2 aims to reduce the number of stunted children 40% by 2025 and eliminate all forms of malnutrition by 2030.

In response to the increasing recognition of malnutrition as a problem of global importance, the Scaling Up Nutrition movement (SUN) was founded in September 2010.^8^ The SUN movement works to end all forms of malnutrition by facilitating coordination between local, regional and international stakeholders.^8^ More than 60 countries have joined the SUN movement by establishing national organizations of nutrition stakeholders that are supported by a high-level individual in the government.^9^ Increasing the coverage of evidence-based nutrition interventions is a central focus of the SUN agenda for improving nutrition in member countries. These interventions are targeted during the “first 1,000-days.” This period of time refers to the critical period of growth and development that occurs between conception and a child’s second birthday.

This study examines child nutrition policy in Guatemala, a Central American country of 16.9 million people. In 2005, Guatemala ratified the National Food and Nutrition Security System (SINASAN in Spanish), establishing a legal framework and political commitment to address stunting.^10^ In 2010, Guatemala joined the SUN movement, affirming many of its previous commitments while incorporating SUN principles into its subsequent national nutrition plans.^11–13^ In 2015, the SUN movement singled out Guatemala as a model case of a country that had made significant progress on addressing nutrition.^14^ In 2017, Guatemala was ranked number one of 45 countries in its political commitment to nutrition in the Hunger and Nutrition Commitment Index.^15^ However, despite more than a decade of political commitment to nutrition, Guatemala still has one of the world’s highest levels of stunting.^2,16^ Overall, 47% of Guatemalan children under the age of 5 years are stunted.^17^ Within Guatemala, there are marked disparities in stunting by wealth status, educational level, and ethnicity that have not substantially improved in the last two decades.^16,18^ Nearly 70% of poor indigenous children in Guatemala are stunted.^19^

In February 2020, the new Guatemalan government led by President Alejandro Giammattei released the most recent national nutrition policy, the *Gran Cruzada Nacional por la Nutrición* (“Great National Crusade for Nutrition,” henceforth “Great Crusade”).^13^ Like previous national nutrition policies that were influenced by the SUN framework, the Great Crusade focuses on evidence-based interventions delivered during the 1,000 days window.^11,12^ Interventions in the Great Crusade include both nutrition-specific and nutrition-sensitive interventions. Nutrition-specific interventions address the immediate determinants of stunting through micronutrient supplementation, breastfeeding promotion, complementary feeding, and other strategies.^20^ Nutrition-sensitive interventions address the structural underpinnings of chronic malnutrition by targeting poverty, education, women’s empowerment, environmental protection, social safety nets, and other targets.^21^ One of the Great Crusade’s principal goals is to reduce the prevalence of child stunting 7% by 2024.^13^

However, it is uncertain if the Great Crusade can reach this ambitious goal. Previous national policies have not met stunting prevalence targets.^22^ Nutrition-specific interventions form the backbone of the Great Crusade’s recommendations, but these interventions tend to have small effect sizes on stunting in clinical trials.^23^ Nutrition-sensitive interventions put forth by the Great Crusade, while promising, have limited evidence of benefit on child growth.^21^ Finally, implementation of nutrition interventions in Guatemala historically has been inconsistent, which may limit the impact of national policies.^24^

The objective of this study was to project the impact of the 2020 Guatemalan national nutrition policy, the Great Crusade. We used a popular maternal and child health modeling program, the Lives Saved Tool (LiST), to carry out our projections. Realistic projections can show the potential impact and uncertainties of nutrition policy in Guatemala and assist in the policy’s implementation. Finally, as a case study in the real-time application of a policy modeling tool, this study may assist policymakers in other low- and middle-income countries who wish to project the impact and uncertainty of maternal and child health policies in their own countries.

## METHODS

This study used LiST to project the policy implementation of the Great Crusade. LiST is a publicly available maternal and child health modeling tool within the Spectrum software package (Avenir Health, Glastonbury, CT, USA). LiST uses publicly available data to project maternal and child outcomes including stunting.^25,26^ LiST was developed in 2003 and is maintained by the Johns Hopkins Bloomberg School of Public Health with funding by the Bill and Melinda Gates Foundation.^27^ Since its development, LiST has been used in more than 110 peer-reviewed research publications.^28^ Many LiST publications have been seminal contributions in the field of global maternal and child health such as *Disease Control Priorities 3rd Edition*^29^ and the 2008 and the 2013 *Lancet* Maternal and Child Undernutrition Series papers.^23^

### Data sources

Data inputs necessary to project stunting in LiST cover four broad categories: (1) demographics, (2) baseline child and maternal health characteristics, (3) intervention coverage levels, and (4) intervention effectiveness.

National-level default input data for most countries including Guatemala are available for download through Spectrum. The default data sources include demographic surveys, academic research, and estimates produced by international bodies such as WHO, UNICEF, UNFPA, World Bank, and United Nations. Default data is updated either annually or as often as national surveys are released.^30^

We carefully reviewed each default input for Guatemala, and, using our knowledge of local data sources, updated inputs with more recent or appropriate values. With one exception, data on intervention effectiveness were not changed from the default values estimated from systematic reviews, meta-analyses, Delphi methods, and randomized control trials.^30^ In May 2020, after consulting with a member of the LiST team at the Johns Hopkins Bloomberg School of Public Health, we were informed of new intervention effectiveness estimates for water, sanitation, and hygiene (WASH) interventions. We then manually updated these estimates in the model. A complete list of data sources and input parameters used in this study can be found in the Appendix.

### Outcomes

Our primary outcomes were changes in stunting prevalence, number of stunting cases averted, and number of stunting cases averted by intervention. Stunting is defined in LiST as a child’s height-for-age more than two standard deviations below the WHO Child Growth Standards median, Z-score (HAZ) < -2.^31^

### Intervention variables

We first reviewed the interventions that impact child stunting in LiST and cross-referenced them with the interventions recommended in the Great Crusade. Of the 15 stunting-related interventions included in LiST, 14 were proposed in the Great Crusade. We subsequently excluded two interventions described in the Great Crusade that are not epidemiologically significant in Guatemala and two additional interventions for which data were not available. Our final model included 10 interventions (Table 1 and Appendix).

**Table 1:**
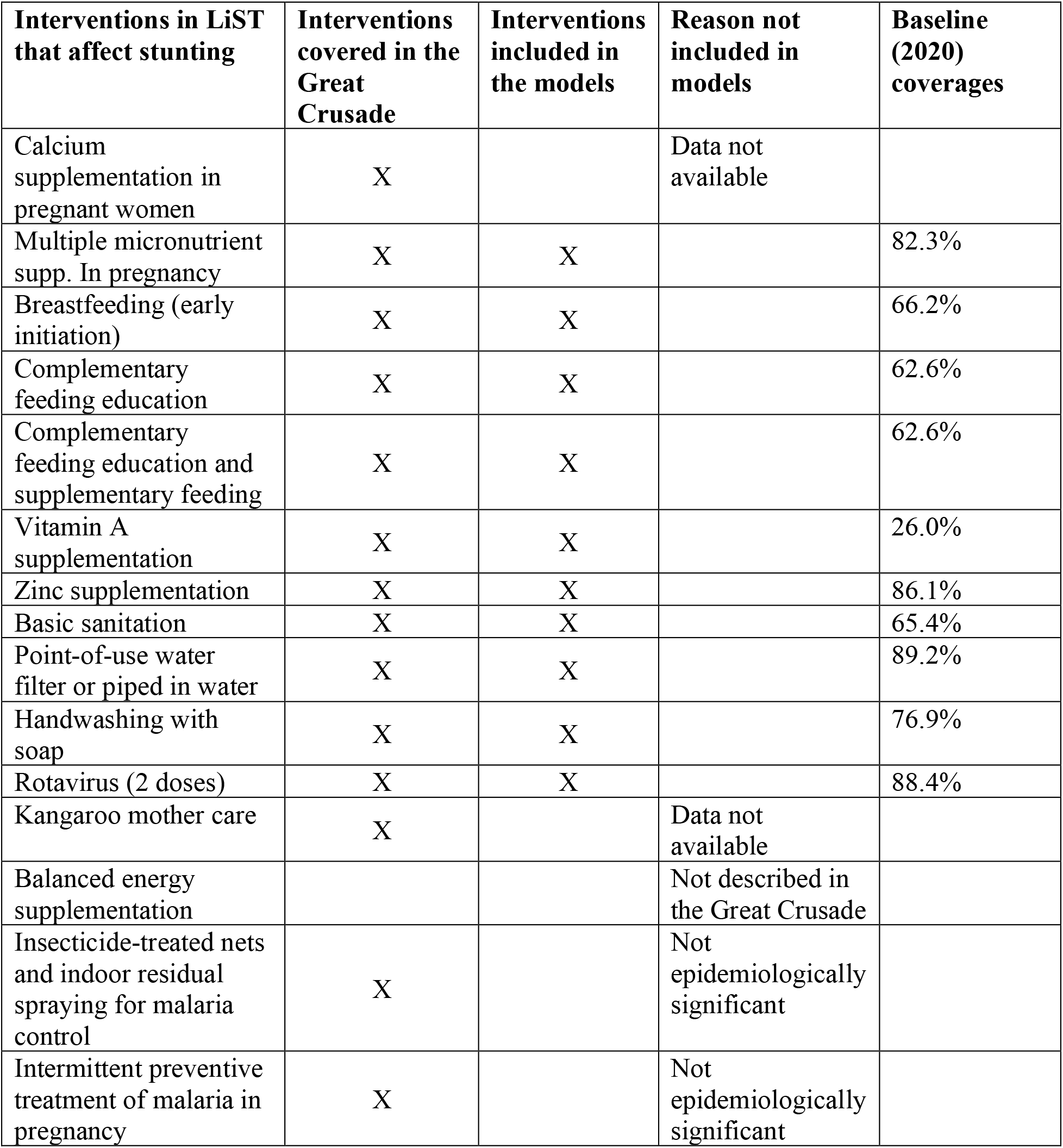
Stunting-related interventions included in the models.

### Scenarios

We modeled four scenarios of intervention coverage change from 2020 to 2030. We chose this period of time because it encompasses both the Great Crusade’s targets in 2024 as well as the Sustainable Development Goal targets in 2030. Each scenario started with the same inputs for the baseline year of 2020 and continued over time based upon the below assumptions. Baseline, 2024, and 2030 coverage levels can be found in the Appendix.

#### Scenario 1: Historical Trends

Our baseline scenario projected future coverage levels based on past historical trends. As described below and similar to prior LiST analyses,^32^ we projected future trends in intervention coverage based on regression models of historical survey data. The key assumption of this scenario is that past intervention coverage trends predict future coverage.

#### Scenario 2: Great Crusade

Intervention coverage levels were set to the goals detailed in the Great Crusade.^13^ From 2024-2030, coverage levels remained static.

#### Scenario 3: Great Crusade with decreased fertility risk

Intervention coverage levels were set to the goals detailed in the Great Crusade. Fertility risk was lowered by eliminating pregnancies before 18 years of age and birth intervals less than 24 months by 2030. This scenario assumed a hypothetical implementation of policies and interventions to eliminate teen pregnancy and short interpregnancy intervals. These measures have been associated with improved child stunting in international and national surveys but were not otherwise included as an intervention in the models.^33,34^

#### Scenario 4: Aspirational coverage

Intervention coverage levels were set to reach 90% by 2024. From 2024-2030, coverage levels remained static.

### Analyses

#### Data preparation

Given that the majority of survey data available in Guatemala was at least three years old, we updated model input parameters for the baseline year of 2020. We projected baseline levels by fitting a logistic regression curve fixed to pass through the last available data point of population-averaged survey data. Shifting the trendline in this manner has been done in a prior LiST study and reflects our greater confidence in more recent survey estimates in Guatemala.^32^ Stata version 16.1 was used in these analyses.

Consistent with previous LiST studies, increases in intervention coverage over time were assumed to increase linearly from the base year.^35,36^ In instances where coverage inputs exceeded 90%, we fixed values at 90% for the remaining years to reflect a reasonable upper bound for coverage. We did not fix the upper bound of WASH interventions, as we believe that these interventions will follow a meaningful trend of improvement beyond this level.

#### LiST model

Previous publications have detailed LiST’s methodology.^27,37,38^ LiST is a linear and deterministic model that uses publicly available data to project maternal and child outcomes including stunting.^25,26^ LiST is implemented within the Spectrum software package (Avenir Health, Glastonbury, CT, USA). Although not a probabilistic model, LiST contains an uncertainty analysis. LiST can assess uncertainty by randomly sampling distributions around the model’s inputs.^39^ We used LiST’s uncertainty analysis tool by running 250 iterations with plausibility bounds set to 95%. Our projections are included as supplementary files. All LiST analyses were done using Spectrum version 5.761.

### Ethics and preregistration

This study uses de-identified data and did not require approval by the Institutional Review Board of the University of Michigan (UM00177760). We preregistered our analysis at the Open Science Foundation on April 13, 2020.^40^

## RESULTS

### Changes in stunting prevalence

The projected impact of scaling up intervention coverage for each scenario is depicted in Figure 1.

**Figure 1:**
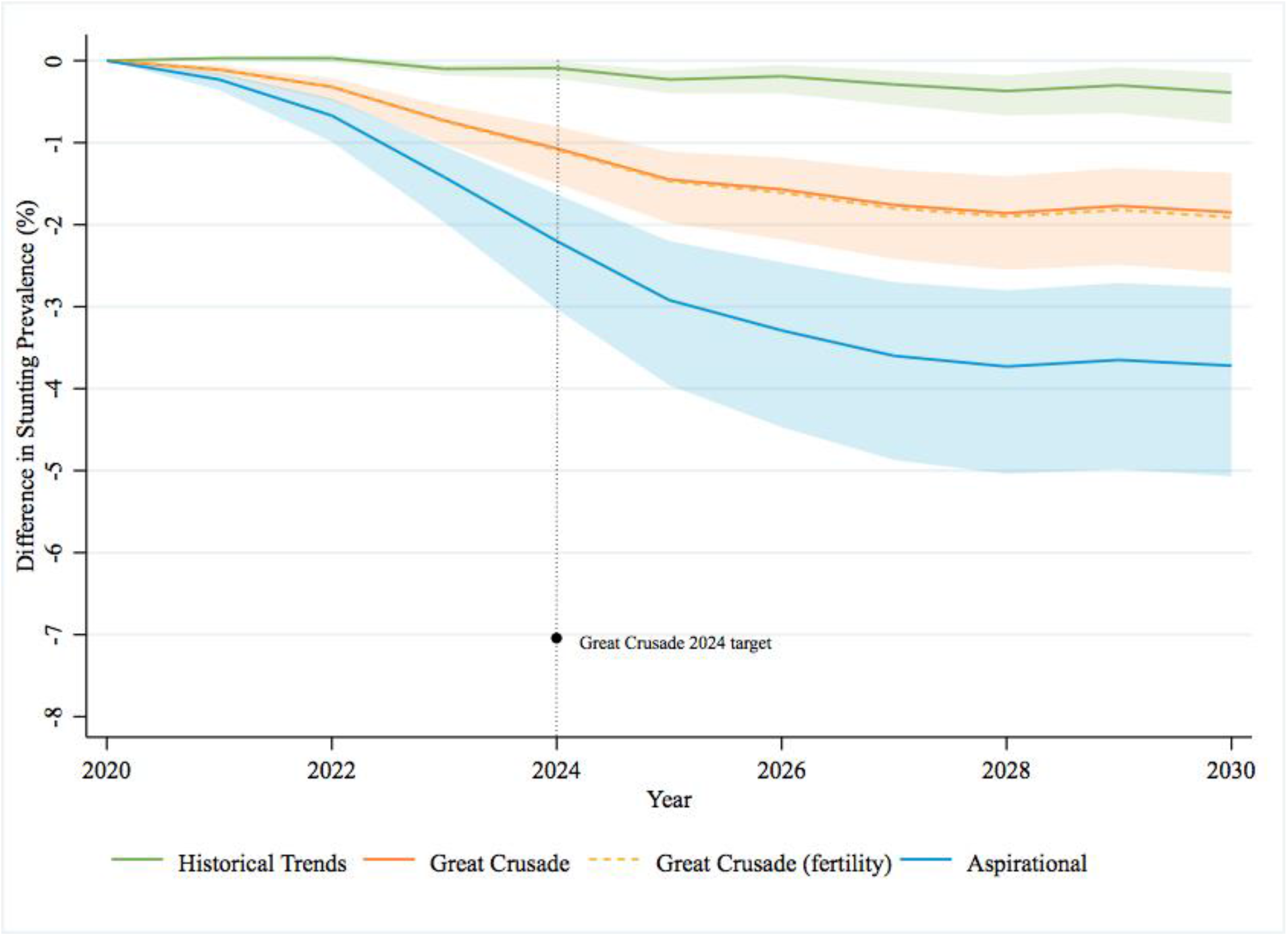
Projected change in stunting prevalence in children under 5 years from the baseline year by scenario. Shaded area represents 95% confidence intervals. Confidence intervals for the Great Crusade (fertility) model are omitted to simplify the figure given overlapping uncertainty bands.

#### 2020-2024 outcomes

If intervention coverage levels increase at historical levels stunting prevalence is projected to change in 2024 by -0.1% (95% confidence interval [CI] 0.0% to -0.2%). The projected change in stunting prevalence in the Great Crusade model by 2024 is a -1.1% (95% CI -0.8% to -1.5%) change. The projected change in stunting prevalence in the Great Crusade with reduced fertility risk model is nearly identical, -1.1% (95% CI -0.8% to -1.6%). Our aspirational model projects a -2.2% (95% CI -1.6% to -3.0%) change in stunting prevalence by 2024.

#### 2020-2030 outcomes

In the Great Crusade model that assumes coverage goals are attained in 2024 and then maintained from 2024-2030, the model projected a change in the prevalence of stunting of -1.8% (95% CI -1.4% to -2.6%). The historical trends and aspirational scenarios projected a change in stunting from 2020-2030 of -0.4% (95% CI -0.2% to -0.8%) and -3.7% (95% CI -2.8% to - 5.1%), respectively.

### Stunting cases averted in 2024 and 2030

Figure 2 shows the estimated number of stunted cases that would be averted by scenario per year.

**Figure 2:**
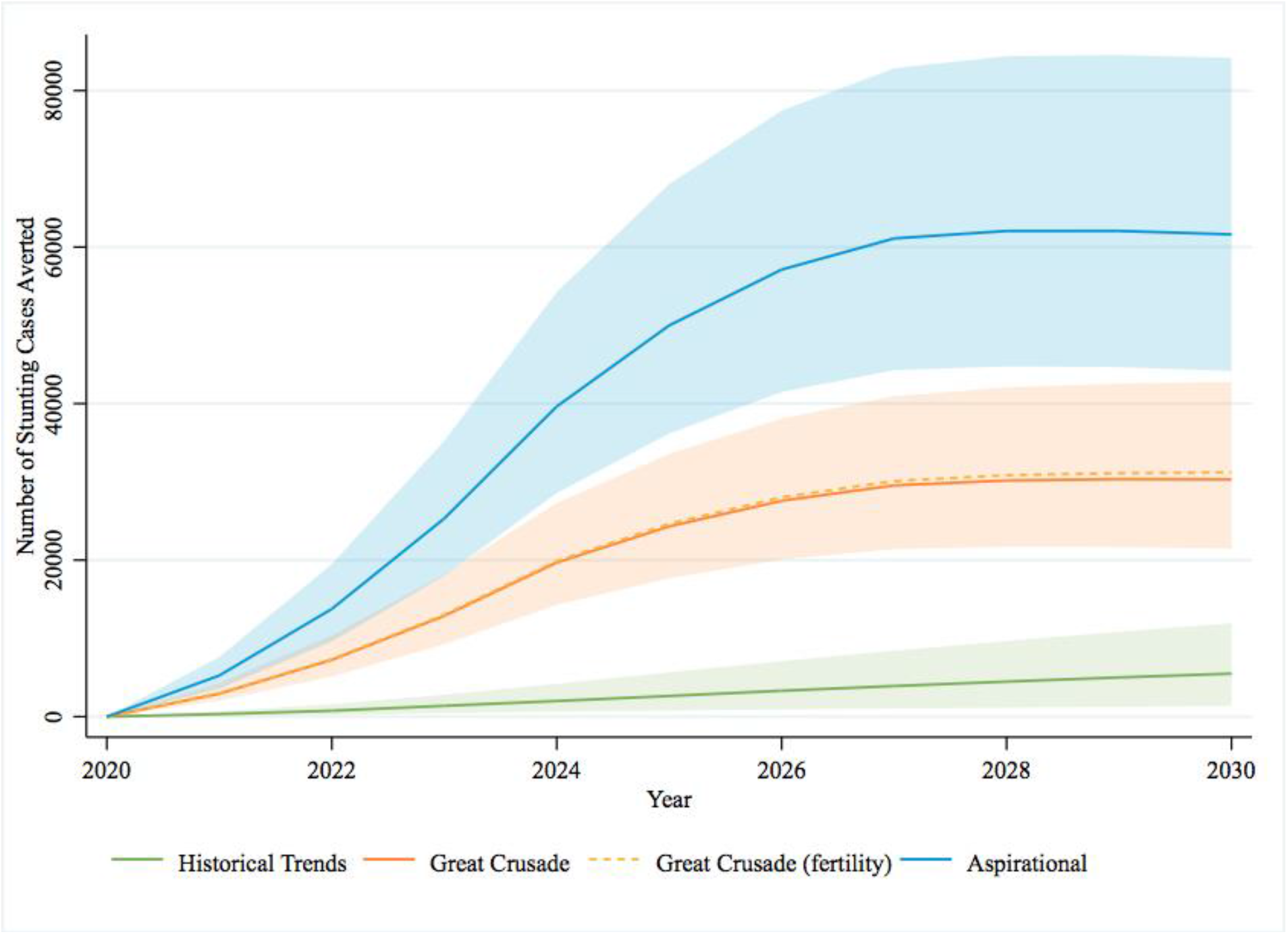
Projected number of stunting cases averted in children under 5 years per year from the baseline year by scenario. Shaded area represents 95% confidence intervals. Confidence intervals for the Great Crusade (fertility) model are omitted to simplify the figure given overlapping uncertainty bands.

#### 2020-2024 outcomes

The estimated number of cumulative stunting cases averted in the historical trends model from 2020-2024 is 4,463. The Great Crusade is projected to avert 42,754 total cases from 2020-2024, and implementation of the Great Crusade along with decreasing fertility risk would avert 43,208 total cases over this time period. The aspirational model is estimated to avert 83,970 total cases of stunting from 2020 to 2024.

#### 2020-2030 outcomes

From 2020-2030, if intervention coverage continues to rise based on historical trends, we have estimated that 29,307 total cases of stunting will be averted. The Great Crusade plan is projected to avert 215,033 total cases during this time, and the Great Crusade model along with decreasing fertility risk would avert 219,167 total cases. The aspirational model is estimated to avert 437,958 total cases of stunting from 2020-2030.

### Stunting cases averted by intervention

Figure 3 elaborates the contribution of each intervention to the cumulative number of stunting cases averted from 2020-2024 (panel A) and 2020-2030 (panel B). In the historical trends model, changes in coverage of point-of-use water filter or piped water primarily contribute to the number of stunting cases averted. Differences in stunting cases averted between the historical trends and the Great Crusade models are driven largely by differential coverage of complementary feeding and sanitation. Differences between the Great Crusade models and aspirational model are largely driven by differential coverage in complementary feeding and breastfeeding promotion.

**Figure 3:**
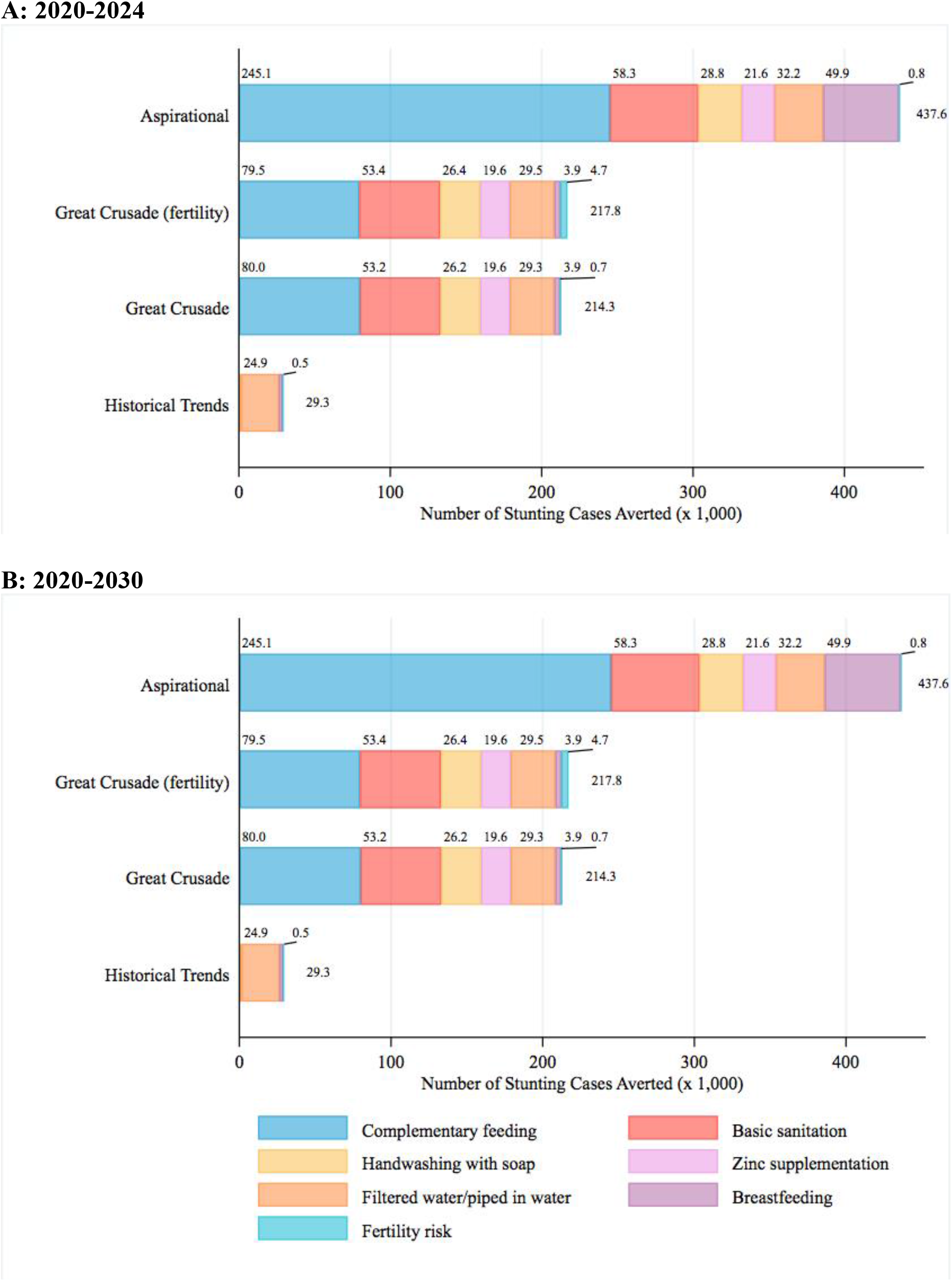
Contribution of interventions on cumulative stunting cases averted by scenario. Only interventions that contribute at least 1.0% of the total are included in colored subsegments.

Figure 4 shows the cumulative contribution of each intervention on stunting cases averted in the aspirational model from 2020 to 2030. This model projected that 56% (245,129 of 437,579) of averted stunting cases would be attributable to increasing coverage of complementary feeding.

**Figure 4:**
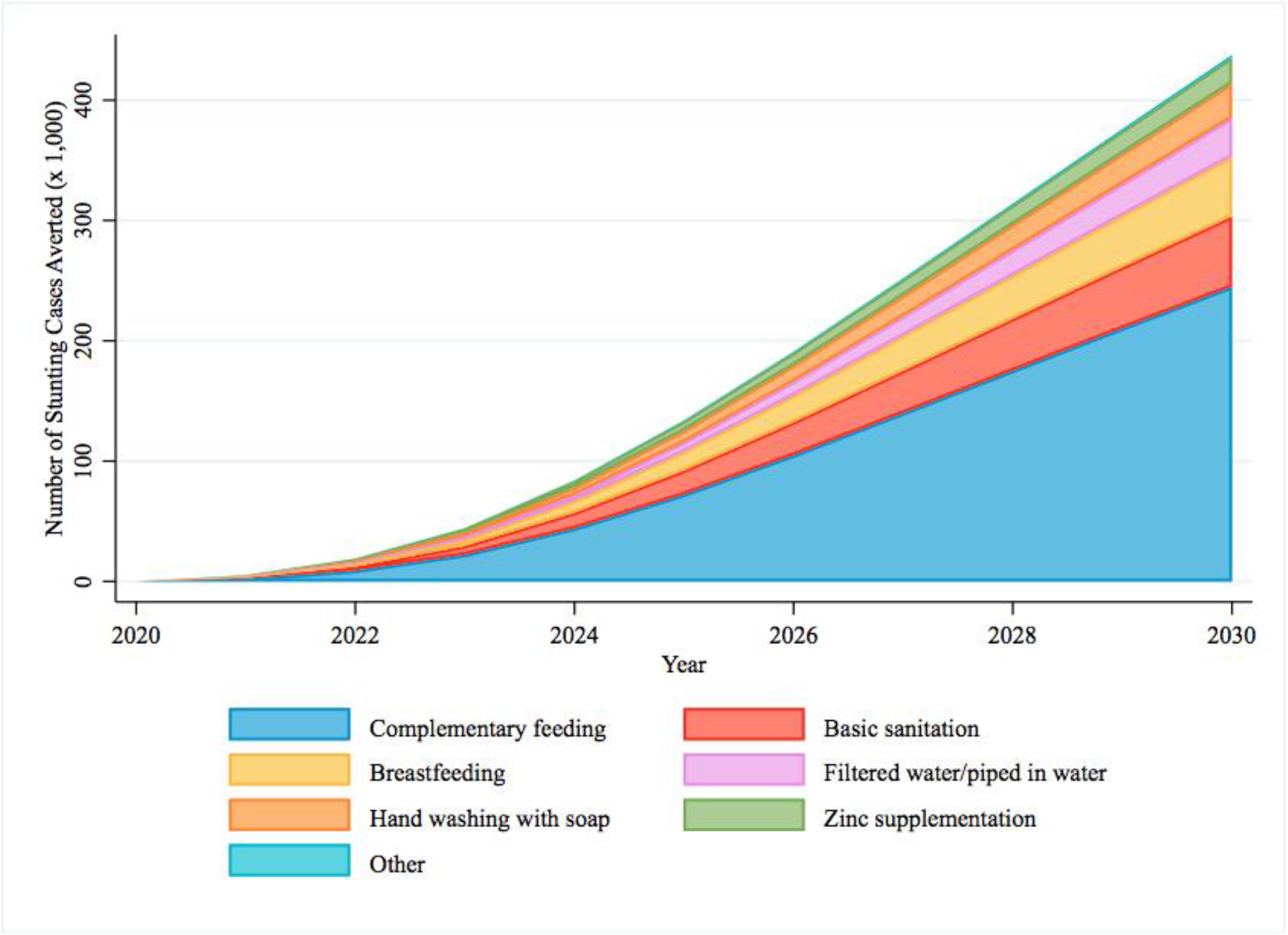
Contribution of interventions on cumulative stunting cases averted in the aspirational model. “Other” category includes multiple micronutrient supplementation, fertility risk, and rotavirus vaccination.

We include the complete output data of our models in the Appendix.

## DISCUSSION

This study used the Lives Saved Tool to project the impact of the 2020 Guatemalan national nutrition policy, the Great Crusade. We found that increases in intervention coverage proposed in the Great Crusade are unlikely to improve child stunting outcomes to a sufficient degree to meet 2024 national targets or 2030 Sustainable Development Goals targets. We also described the uncertainty of our projections and the relative impact of different interventions. Our study has implications for the optimization of nutrition in Guatemala and other low- and middle-income countries.

Our modeling study of nutrition policy in Guatemala reveals limitations in the SUN framework that focuses on scaling-up evidenced based nutrition interventions. Despite strong political commitment to the SUN movement since 2010, Guatemala has not met historical stunting targets and, as our study shows, may have difficulty meeting future goals. In the 2013 Lancet series on Maternal and Child Health, it was estimated that increasing the coverage of ten evidence-based interventions to 90% in 34 high burden countries would result in a 20% relative reduction in stunting prevalence.^23^ Our models suggest that increasing intervention coverage to 90% in Guatemala would result in an 4.7% relative reduction in stunting in 2024 and 8.0% relative reduction in 2030. The modest impact of nutrition coverage expansion in Guatemala compared with other countries may be explained by the relatively high baseline intervention coverage in Guatemala. Taken together, our findings imply that there may be substantial heterogeneity in impact between countries that implement evidence-based nutrition stunting interventions endorsed within the SUN framework. In other country cases studies, contextual differences and variations in investments both within and outside the health sector have been proposed as explanatory factors for improvements in stunting.^41^

Our findings also reveal the limitations of currently available evidence to prevent stunting. Except for WASH, all of the stunting interventions incorporated in LiST and included in our models are nutrition-specific interventions. These interventions are supported by high-level evidence, but, in general, their effect sizes are modest. As one example, complementary food supplementation in food-insecure populations causes a 0.10 (95% CI: 0.03 to 0.17) improvement in length-for-age Z-scores.^42^ Nutrition-sensitive interventions focusing on broader context of nutrition including poverty, agriculture, social safety nets, education, and other areas as have been proposed as a potential way to generate more sizeable reductions in stunting.^43,44^ In Peru, for example, large improvements in child stunting observed from 2000-2013 were likely attributable to economic growth, increased societal participation, poverty-reduction strategies, and increased health spending.^45,46^ Furthermore, a retrospective review of five exemplar countries that made significant improvements in stunting found that nutrition-sensitive strategies accounted for 50% of the total stunting reduction.^41^ Despite compelling ecological evidence, nutrition-sensitive interventions have not consistently shown improvements in linear growth in randomized controlled trials. Future research is urgently needed in this area.^21^

Our study supports the emerging criticism of SUN’s emphasis on technical solutions to address stunting. An independent review of the SUN movement has acknowledged that: (1) there is limited evidence that SUN has improved nutrition outcomes of member countries, (2) some countries including Guatemala that closely adhere to SUN have not observed meaningful improvements in stunting, and (3) SUN’s standardized approach does not sufficiently account for local country factors.^47^ Other critics have argued that SUN emphasizes short-term technical solutions with limited consideration of the structural causes of stunting.^48^ In this view, even SUN’s nutrition-sensitive interventions primarily serve to benefit commercial food systems that disrupt indigenous food cultures, decrease confidence in local foods, and reduce biodiversity.^48^

In Guatemala and elsewhere, critics of SUN have pointed out conflicts of interest,^49^ human-rights concerns among adolescent mothers,^50,51^ and detachment from communities affected by malnutrition.^48^ While our study does not address these broader critiques, our results buttress criticisms against SUN’s emphasis on narrow technical interventions and call for increased focus on strategies that address the social determinants that give rise to stunting.

With respect to our study’s implications in Guatemala, a principal target of the Great Crusade is a 7% reduction in stunting prevalence from 2020-2024. Our study suggests that this target is very ambitious and unlikely to be achieved solely based on scaling-up interventions recommended by SUN and outlined in the Great Crusade. According to our projections, even aspirational levels of coverage would only reduce stunting prevalence by -2.2% (95% CI -1.6% to -3.0%) in 2024, - 0.6% per year. Previous reviews of countries that made significant strides in improving stunting have shown that per year reductions in stunting greater than 1% are possible.^45,52^ The previous Guatemalan national nutrition policies from 2012-2016^11^ and 2016-2020^12^ also were unable to reach targets of a 10% absolute reduction in stunting. Importantly, while the Great Crusade confers only a small absolute reduction in stunting prevalence in our models, we also project that the policy would avert over 42,000 total stunting cases from 2020-2024 and 217,000 total cases from 2020-2030. This absolute number of cases averted may be viewed as substantial by policymakers and nutrition stakeholders in a country with a total population of 16.9 million people.

We found dramatic differences in impact among interventions recommended by SUN and included in the Great Crusade. Complementary feeding and basic sanitation contribute to more than half of the stunting cases averted in most of our models. Complementary feeding has a particularly large impact in the aspirational models, and we estimate that increasing in complementary feeding intervention coverage to 90% by 2024 would avert 245,000 total stunting cases from 2020-2030. After complementary feeding, the intervention contributing the greatest impact on stunting reduction between the Great Crusade and aspirational models was breastfeeding promotion. Interventions such as complementary feeding and breastfeeding promotion are “double-duty actions” that also may confer beneficial impacts on obesity and other diet-related non-communicable diseases.^53^ Notably, some interventions such as multiple micronutrient supplementation in pregnancy, rotavirus vaccine, and vitamin A supplementation had minimal impact on stunting due to high levels of coverage at baseline, small effect sizes or limited need. Previous research has suggested an association between family planning and child stunting,^33^ but our models found that decreasing fertility risks had a very small impact on stunting. Our results suggest that efforts to reduce stunting should focus resources on scaling-up coverage of specific interventions (complementary feeding, basic sanitation, and breastfeeding) over others (multiple micronutrient supplementation in pregnancy, rotavirus vaccine, and vitamin A supplementation) during the Great Crusade’s implementation phase.

In addition to our evaluation of nutrition in Guatemala, our study makes a useful methodological contribution to the broader literature that uses LiST and other modeling tools to project maternal and child health outcomes. To our knowledge, this is the first study that (1) uses LiST to project the impact of a national health policy as the policy is launched or (2) assesses the uncertainty of LiST estimates. Policy analyses often report exact predictions without assessments of uncertainty, a practice criticized in the literature as “incredible certitude.”^54^ We agree that uncertainty should be transparently and appropriately communicated in policy analyses including maternal and child health models such as LiST.^55^ We depict the uncertainty of our models in Figures 2 and 3 where greater changes in intervention coverage and time are accompanied by larger uncertainty.

Strengths of this study include our choice of a well-established modeling tool (LiST), and our detailed assessments of uncertainty. We also updated default inputs with new and non-public data sources. Finally, we addressed the gap between the last Demographic and Health Survey in Guatemala (2014-2015) and the first year of our projection (2020) by estimating baseline 2020 inputs using a methodology from the literature.

Our study has several limitations. First, our study assesses only a single country, Guatemala. We justify our focus on Guatemala given the country’s unique position of extraordinary stunting prevalence and sustained commitment to SUN. Furthermore, country case studies have a rich tradition in informing global policy within the maternal and child health field.^45^ Second, LiST was designed to estimate the impact of evidence-based interventions with known effect sizes. We are unable to estimate the impact of nutrition-sensitive interventions in the Great Crusade that have uncertain effect sizes such as education and conditional-cash transfer programs. Third, our study focuses on linear growth—represented by stunting—as a central metric of early child health. However, there has been increasing criticism of the causal assumptions between linear growth in children and long-term developmental outcomes.^56^ Pioneering studies in Jamaica^57–59^ and meta-analyses^60,61^ show that early childhood education and stimulation interventions can improve child development without improving growth. In Guatemala, community based interventions targeting the determinants of child development (and not solely growth) are important but are not considered in our study.^62,63^

A final limitation involves the uncertainty of nutrition policy in Guatemala during the COVID-19 pandemic. The Great Crusade was published in February 2020 before the first documented case of COVID-19 in Guatemala. Undoubtedly, COVID-19 will have direct and indirect effects on nutrition in Guatemala. Reports of increased food security and acute malnutrition have emerged,^64,65^ while funds destined for nutrition programs are being rerouted to fight COVID-19.^66^ We chose not to consider COVID-19 in our analysis for three reasons: (1) our primary goal was to assess the Great Crusade as a policy document, (2) the lack of currently available COVID-19 data in Guatemala make any long-term projections impractical, and (3) modeling health system shocks in LiST requires different modeling techniques and assumptions.^67^

## CONCLUSION

This modeling study using LiST found that that the most recent Guatemalan national nutrition policy, the Great Crusade, will have difficulty achieving 2024 national targets or 2030 Sustainable Development Goals targets solely based on increases in intervention coverage. Our results show the limitations of technical solutions to nutrition put forth by the international nutrition community. Despite more than a decade of political commitment on the issue of nutrition, Guatemala may continue to have exceptionally high prevalence of stunting over the coming decade. We recommend the prioritization of interventions that are projected to confer the largest impact such as complementary feeding, breastfeeding, and basic sanitation. Policies and strategies are needed that address the broader social structures that predispose children to stunting.

## Data Availability

Except for one survey, all of our data is from publicly available data sets. We include in the Appendix all of the data inputs. We also include our modeling files as supplementary files.

https://doi.org/10.7910/DVN/LXJVDV

