## Appendices for "Projecting the impact of nutrition policy to improve child stunting: A case study in Guatemala using the Lives Saved Tool"

**INTERVENTIONS INCLUDED IN THE MODEL**

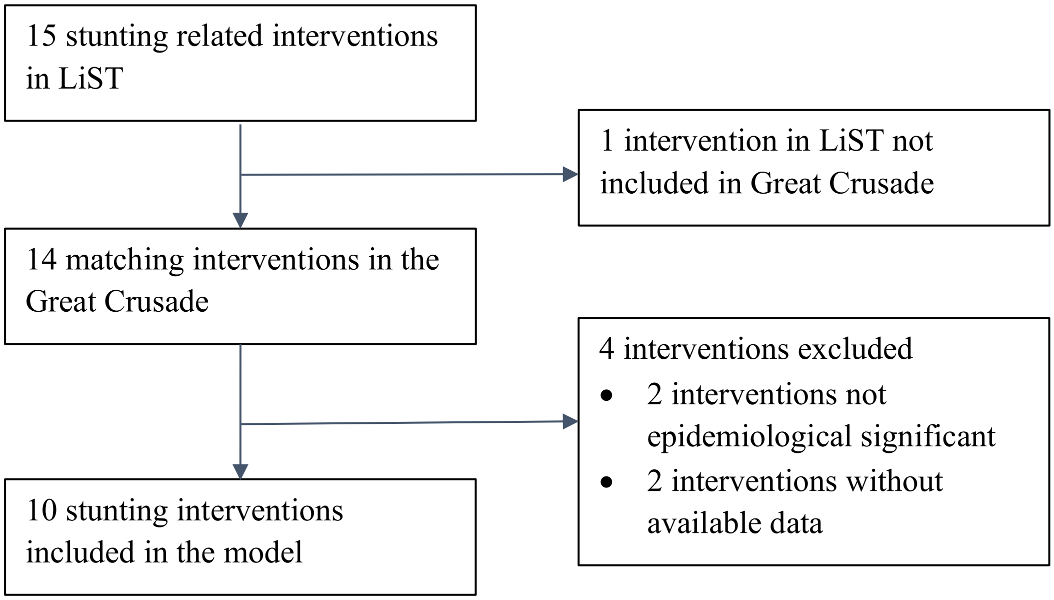

#### MODEL INPUTS

##### Demographics

*First year population single age*

| Age | Male | Female | | Age | Male | Female |
|---|---|---|---|---|---|---|
| **0** | 193,140 | 184,360 | | **41** | 87,311 | 97,985 |
| **1** | 193,084 | 184,827 | | **42** | 83,725 | 95,009 |
| **2** | 193,486 | 185,541 | | **43** | 80,193 | 92,056 |
| **3** | 192,847 | 184,749 | | **44** | 76,729 | 89,056 |
| **4** | 191,977 | 183,962 | | **45** | 73,357 | 85,974 |
| **5** | 191,156 | 183,246 | | **46** | 70,112 | 82,802 |
| **6** | 190,401 | 182,608 | | **47** | 66,987 | 79,328 |
| **7** | 188,802 | 181,171 | | **48** | 63,960 | 75,996 |
| **8** | 186,540 | 179,120 | | **49** | 61,013 | 72,636 |
| **9** | 185,852 | 178,580 | | **50** | 58,159 | 69,338 |
| **10** | 183,284 | 176,249 | | **51** | 55,397 | 66,175 |
| **11** | 182,564 | 175,701 | | **52** | 52,767 | 63,202 |
| **12** | 181,812 | 175,138 | | **53** | 50,260 | 60,413 |
| **13** | 181,017 | 174,554 | | **54** | 47,895 | 57,801 |
| **14** | 180,174 | 173,973 | | **55** | 45,699 | 55,357 |
| **15** | 179,270 | 173,387 | | **56** | 43,737 | 53,440 |
| **16** | 178,264 | 172,757 | | **57** | 41,949 | 51,298 |
| **17** | 177,084 | 172,004 | | **58** | 40,261 | 49,220 |
| **18** | 182,554 | 177,758 | | **59** | 38,640 | 47,185 |
| **19** | 185,290 | 180,995 | | **60** | 37,047 | 45,184 |
| **20** | 183,174 | 179,629 | | **61** | 35,475 | 43,235 |
| **21** | 175,274 | 172,717 | | **62** | 33,937 | 41,329 |
| **22** | 170,249 | 168,605 | | **63** | 32,441 | 39,478 |
| **23** | 165,229 | 164,481 | | **64** | 30,997 | 37,685 |
| **24** | 162,470 | 162,513 | | **65** | 29,603 | 35,944 |
| **25** | 159,398 | 160,214 | | **66** | 28,280 | 34,255 |
| **26** | 155,974 | 157,453 | | **67** | 27,022 | 32,624 |
| **27** | 152,121 | 154,213 | | **68** | 25,821 | 31,032 |
| **28** | 147,939 | 150,618 | | **69** | 24,654 | 29,469 |
| **29** | 143,401 | 146,664 | | **70** | 23,426 | 27,881 |
| **30** | 138,517 | 142,395 | | **71** | 22,346 | 26,413 |
| **31** | 133,356 | 137,920 | | **72** | 21,259 | 24,963 |
| **32** | 128,035 | 133,330 | | **73** | 20,142 | 23,507 |
| **33** | 122,667 | 128,704 | | **74** | 18,985 | 22,028 |
| **34** | 117,383 | 124,132 | | **75** | 17,793 | 20,531 |
| **35** | 112,281 | 119,709 | | **76** | 16,569 | 19,046 |
| **36** | 107,450 | 115,479 | | **77** | 15,346 | 17,601 |
| **37** | 102,928 | 111,485 | | **78** | 14,133 | 16,212 |
| **38** | 98,734 | 107,772 | | **79** | 12,943 | 14,884 |
| **39** | 94,718 | 104,258 | | **80+** | 90,535 | 108,890 |

| | | | | | | |
|---|---|---|---|---|---|---|
| **40** | 90,962 | 101,046 | | **Total** | 8,297,763 | 8,560,479 |

Source: Estimaciones *y Proyecciones Nacionales de Población Metodología y Principales Resultados*. Guatemala City; 2019. (2019 Guatemalan Census)

*Total fertility rate*

| | 2020 | 2021 | 2022 | 2023 | 2024 | 2025 | 2026 | 2027 | 2028 | 2029 | 2030 |
|---|---|---|---|---|---|---|---|---|---|---|---|
| **TFR** | 2.6 | 2.5 | 2.4 | 2.4 | 2.3 | 2.3 | 2.2 | 2.2 | 2.2 | 2.1 | 2.1 |

Source: 2019 Guatemalan census

*Age-specific fertility rate*

| **Age** | **2020** | **2021** | **2022** | **2023** | **2024** | **2025** | **2026** | **2027** | **2028** | **2029** | **2030** |
|---|---|---|---|---|---|---|---|---|---|---|---|
| **15-19** | 12.17 | 12.12 | 12.08 | 12.02 | 11.90 | 11.90 | 11.83 | 11.76 | 11.68 | 11.60 | 11.51 |
| **20-24** | 24.27 | 24.24 | 24.21 | 24.18 | 24.10 | 24.10 | 24.06 | 24.01 | 23.96 | 23.90 | 23.85 |
| **25-29** | 23.55 | 23.58 | 23.61 | 23.64 | 23.73 | 23.73 | 23.77 | 23.82 | 23.87 | 23.93 | 23.99 |
| **30-34** | 19.38 | 19.42 | 19.46 | 19.52 | 19.64 | 19.64 | 19.71 | 19.78 | 19.87 | 19.96 | 20.05 |
| **35-39** | 13.08 | 13.09 | 13.10 | 13.12 | 13.16 | 13.16 | 13.18 | 13.21 | 13.23 | 13.26 | 13.28 |
| **40-44** | 6.10 | 6.09 | 6.08 | 6.07 | 6.05 | 6.05 | 6.03 | 6.02 | 6.00 | 5.99 | 5.97 |
| **45-49** | 1.47 | 1.46 | 1.45 | 1.44 | 1.42 | 1.42 | 1.41 | 1.40 | 1.38 | 1.37 | 1.36 |
| **Total** | 100.02 | 100 | 99.99 | 99.99 | 100 | 100 | 99.99 | 100 | 99.99 | 100.01 | 100.01 |

Source: Default data, World Population Prospects - Population Division - United Nations. https://population.un.org/wpp/

*Sex ratio at birth*

| | 2020 | 2021 | 2022 | 2023 | 2024 | 2025 | 2026 | 2027 | 2028 | 2029 | 2030 |
|---|---|---|---|---|---|---|---|---|---|---|---|
| **Birth ratio** | 103.7 | 103.7 | 103.7 | 103.7 | 103.7 | 103.7 | 103.7 | 103.7 | 103.7 | 103.7 | 103.7 |

Source: Calculated by authors using microdata from: International Ministerio de Salud Pública y Asistencia Social (MSPAS), Instituto Nacional de Estadística (INE), ICF International. *Encuesta Nacional de Salud Materno Infantil 2014-2015. Informe Final*.; 2017. (ENSMI)

*Life expectancy*

| | 2020 | 2021 | 2022 | 2023 | 2024 | 2025 | 2026 | 2027 | 2028 | 2029 | 2030 |
|---|---|---|---|---|---|---|---|---|---|---|---|
| **Male** | 69.9 | 70.1 | 70.3 | 70.5 | 70.7 | 70.8 | 71 | 71.2 | 71.4 | 71.6 | 71.8 |
| **Female** | 76.4 | 76.6 | 76.8 | 77 | 77.1 | 77.3 | 77.5 | 77.7 | 77.9 | 78 | 78.2 |

Source: 2019 Guatemalan census

*International migration*

| | 2020 | 2021 | 2022 | 2023 | 2024 |
|---|---|---|---|---|---|
| **Male** | -21,098.20 | -19,643.90 | -18,346.30 | -17,116.30 | -16,030.90 |
| **Female** | -17,876.60 | -17,380.20 | -16,827.10 | -16,296.50 | -15,711.40 |

| | 2025 | 2026 | 2027 | 2028 | 2029 |
|---|---|---|---|---|---|
| **Male** | -15,153.70 | -14,323.70 | -13,608.60 | -12,928.60 | -12283.6 |
| **Female** | -14,998.10 | -14,317.80 | -13,602.60 | -12,922.40 | -12277.2 |

| | 2030 |
|---|---|
| **Male** | -11668.6 |
| **Female** | -11662.1 |

Source 2019 Guatemalan census

#### Health, Mortality and Economic Status

*Baseline child health status*

Meningococcal A vaccine recommended

[ ] Yes [ X ] No

Nutrition Deficiencies

| Input | Value | Source |
|---|---|---|
| **Percent vitamin A deficient** | 0.0% | SIVESNU 2016 |
| **Percent zinc deficient** | 13.3% | SIVESNU 2016 |

Status at birth (percent of total births)

| Input | Value | Source |
|---|---|---|
| **Preterm: Small for gestational age (PEG)** | 1.61% | Default data<br><br>Lee AC, Katz J, Blencowe H, et al. National and regional estimates of term and preterm babies born small for gestational age in 138 low-income and middle-income countries in 2010. Lancet Global Health 2013; 1(1): e26-36. http://www.ncbi.nlm.nih.gov/pubmed/25103583. |
| **Preterm: Appropriate for gestational age (AEG)** | 6.10% | |
| **Term: Small for gestational age (PEG)** | 11.96% | |
| **Term: Appropriate for gestational age (AEG)** | 80.33% | |
| **Total** | 100% | |

Incidence (Number of cases per child per year)

| | **< 1 month** | **1-5 months** | **6-11 months** | **12-23 months** | **24-59 months** | **Source** |
|---|---|---|---|---|---|---|
| **Incidence of diarrhea** | 3.4 | 3.2 | 3.2 | 3.2 | 3.2 | Default data<br><br>Fischer Walker CL, Rudan I, Liu L, et al. Global burden of childhood pneumonia and diarrhoea. The Lancet 2013; 381(9875): 1405-16. http://www.ncbi.nlm.nih.gov/pubmed/23582727. |
| **Incidence of severe diarrhea** | 0.062 | 0.062 | 0.062 | 0.062 | 0.062 | |
| **Incidence of severe pneumo-nia** | 0.027 | 0.027 | 0.027 | 0.027 | 0.027 | Default data<br><br>Rudan I, O'Brien KL, Nair H, et al. Epidemiology and etiology of childhood pneumonia in 2010: Estimates of incidence, severe morbidity, mortality, underlying risk factors and causative pathogens for 192 countries. Journal of Global Health 2013; 3(1). http://www.ncbi.nlm.nih.gov/pubmed/23826505. (Online supplementary material.) |

| | | | | | | |
|---|---|---|---|---|---|---|
| **Incidence of meningit-is** | 0.001 | 0.001 | 0.001 | 0.001 | 0.001 | Default data<br><br>Calculated from the following sources:<br><br>Davis S, Feikin D, Johnson HL. The effect of Haemophilus influenzae type B and pneumococcal conjugate vaccines on childhood meningitis mortality: A systematic review. BMC Public Health 2013; 13(Suppl 3): S21. http://www.ncbi.nlm.nih.gov/pubmed/24564188.<br><br>See also the WHO online companion table referenced in the following articles: Watt JP, Wolfson LJ, O'Brien KL, et al. Burden of disease caused by Haemophilus influenzae type b in children younger than 5 years: Global estimates. Lancet 2009; 374(9693): 903-911. http://www.ncbi.nlm.nih.gov/pubmed/19748399.<br><br>O'Brien KL, Wolfson LJ, Watt JP, et al. Burden of disease caused by Streptococcus pneumoniae in children younger than 5 years: Global estimates. Lancet 2009; 374(9693): 893-902. http://www.ncbi.nlm.nih.gov/pubmed/19748398. |

*Nutrition status distributions*

#### Stunting

| | Percent | Source |
|---|---|---|
| **Stunting** | 46.47% | Automatically calculated from stunting distribution. |

| Stunting distribution | <1 month | 1-5 months | 6-11 months | 12-23 months | 24-59 months | Source |
|---|---|---|---|---|---|---|
| **Greater than 1 SD less than the median norm** | 36.03% | 36.03% | 31.34% | 19.07% | 19.84% | Calculated by authors using microdata from ENSMI 2014-2015. |
| **Between 1 and 2 SDs less than the median norm** | 34.01% | 34.01% | 34.24% | 30.16% | 30.37% | |
| **Between 2 and 3 SDs less than the median norm** | 22.49% | 22.49% | 23.94% | 29.07% | 31.94% | |
| **More than 3 standard deviations less than the median norm** | 7.466% | 7.466% | 10.49% | 21.7% | 17.85% | |
| **Total** | 100.00% | 100.00% | 100.01% | 100.00% | 100.00% | |

#### Wasting

| | Percent | Source |
|---|---|---|
| **Wasting** | 0.72% | Automatically calculated from wasting distribution. |

| Wasting distribution | <1 month | 1-5 months | 6-11 months | 12-23 months | 24-59 months | Source |
|---|---|---|---|---|---|---|
| **Greater than 1 SD less than the median norm** | 96.63% | 96.63% | 92.96% | 88.27% | 93.13% | Calculated by authors using microdata from ENSMI 2014-2015. |
| **Between 1 and 2 SDs less than the median norm** | 2.384% | 2.384% | 6.338% | 10.6% | 6.313% | |
| **Between 2 and 3 SDs less than the** | 0.6523% | 0.6523% | 0.6406% | 1.077% | 0.4344% | |

| | | | | | |
|---|---|---|---|---|---|
| **median norm** | | | | | |
| **More than 3 standard deviations less than the median norm** | 0.3345% | 0.3345% | 0.0591% | 0.0516% | 0.1205% |
| **Total** | 100.00% | 100.00% | 100.00% | 100.00% | 100.00% |

*Pathogens*

Diarrhea – distribution among all cases

| | 1-5 months | 6-11 months | 12-23 months | 24-59 months |
|---|---|---|---|---|
| **Rotavirus** | 5.5% | 5.5% | 5.5% | 5.5% |
| **Pathogen B** | 0% | 0% | 0% | 0% |
| **Pathogen C** | 0% | 0% | 0% | 0% |
| **All other pathogens** | 94.5% | 94.5% | 94.5% | 94.5% |
| **Total** | 100% | 100%% | 100% | 100% |

Diarrhea – distribution among severe cases

| | 1-5 months | 6-11 months | 12-23 months | 24-59 months |
|---|---|---|---|---|
| **Rotavirus** | 0% | 0% | 0% | 0% |
| **Pathogen B** | 23.4% | 23.4% | 23.4% | 23.4% |
| **Pathogen C** | 0% | 0% | 0% | 0% |
| **All other pathogens** | 76.6% | 76.6% | 76.6% | 76.6% |
| **Total** | 100% | 100% | 100% | 100% |

Diarrhea – distribution among fatal cases

| | 1-5 months | 6-11 months | 12-23 months | 24-59 months |
|---|---|---|---|---|
| **Rotavirus** | 23.4% | 23.4% | 23.4% | 23.4% |
| **Pathogen B** | 0% | 0% | 0% | 0% |
| **Pathogen C** | 0% | 0% | 0% | 0% |
| **All other pathogens** | 76.6% | 76.6% | 76.6% | 76.6% |
| **Total** | 100% | 100% | 100% | 100% |

Source: Default data, Fischer Walker CL, Rudan I, Liu L, et al. Global burden of childhood pneumonia and diarrhoea. The Lancet 2013; 381(9875): 1405-16. http://www.ncbi.nlm.nih.gov/pubmed/23582727

Pneumonia – distribution among severe cases

| | 1-5 months | 6-11 months | 12-23 months | 24-59 months |
|---|---|---|---|---|
| **H. influenzae type b** | 4% | 4% | 4% | 4% |
| **S. pneumoniae** | 7% | 7% | 7% | 7% |
| **Influenza virus** | 10.7% | 10.7% | 10.7% | 10.7% |
| **All other pathogens** | 78.3% | 78.3% | 78.3% | 78.3% |
| **Total** | 100% | 100% | 100% | 100% |

Pneumonia – distribution among fatal cases

| | 1-5 months | 6-11 months | 12-23 months | 24-59 months |
|---|---|---|---|---|
| **H. influenzae type b** | 21.3% | 21.3% | 21.3% | 21.3% |
| **S. pneumoniae** | 32.8% | 32.8% | 32.8% | 32.8% |

| | | | | |
|---|---|---|---|---|
| **Influenza virus** | 10.7% | 10.7% | 10.7% | 10.7% |
| **All other pathogens** | 35.2% | 35.2% | 35.2% | 35.2% |
| **Total** | 100% | 100% | 100% | 100% |

Source: Default data, Rudan I, O'Brien KL, Nair H, et al. Epidemiology and etiology of childhood pneumonia in 2010: Estimates of incidence, severe morbidity, mortality, underlying risk factors and causative pathogens for 192 countries. Journal of Global Health 2013; 3(1). http://www.ncbi.nlm.nih.gov/pubmed/23826505. Please refer to the online supplementary document

Meningitis – distribution among severe cases

| | **1-5 months** | **6-11 months** | **12-23 months** | **24-59 months** |
|---|---|---|---|---|
| **H. influenzae type b** | 46% | 46% | 46% | 46% |
| **S. pneumoniae** | 52% | 52% | 52% | 52% |
| **N. meningitis type A** | 2% | 2% | 2% | 2% |
| **All other pathogens** | 0% | 0% | 0% | 0% |
| **Total** | 100% | 100% | 100% | 100% |

Meningitis – distribution among fatal cases

| | **1-5 months** | **6-11 months** | **12-23 months** | **24-59 months** |
|---|---|---|---|---|
| **H. influenzae type b** | 46% | 46% | 46% | 46% |
| **S. pneumoniae** | 52% | 52% | 52% | 52% |
| **N. meningitis type A** | 2% | 2% | 2% | 2% |
| **All other pathogens** | 0% | 0% | 0% | 0% |
| **Total** | 100% | 100% | 100% | 100% |

Source: Default data, Davis S, Feikin D, Johnson HL. The effect of Haemophilus influenzae type B and pneumococcal conjugate vaccines on childhood meningitis mortality: A systematic review. BMC Public Health 2013; 13(Suppl 3): S21. http://www.ncbi.nlm.nih.gov/pmc/articles/PMC3847464/

*Baseline child mortality*

#### Mortality rate

| | **Mortality rate** |
|---|---|
| **Neonatal** | 11.58 |
| **Infant** | 20.56 |
| **Child** | 24.19 |

Source: Calculated by authors using historical data from: UN Inter-agency Group for Child Mortality Estimation. CME info - Child Mortality Estimates. http://childmortality.org.

#### Percent of child deaths by proximate cause

| **Neonatal** | **Percent** | **Post neonatal** | **Percent** |
|---|---|---|---|
| **Diarrhea** | 0.4% | **Diarrhea** | 13.1% |
| **Sepsis** | 16.8% | **Pneumonia** | 23.5% |
| **Pneumonia** | 5.9% | **Meningitis** | 1.9% |
| **Asphyxia** | 25.0% | **Measles** | 0.0% |
| **Prematurity** | 27.2% | **Malaria** | 0.0% |
| **Tetanus** | 0.0% | **Pertussis** | 1.0% |
| **Congenital anomalies** | 16.5% | **AIDS** | 1.2% |
| **Other** | 8.3% | **Injury** | 16.7% |
| **Total** | 100.0% | **Other** | 42.6% |
| | | **Total** | 100.0% |

Source: Default data, WHO estimates for years 2000-2017.
https://www.who.int/healthinfo/global_burden_disease/estimates/en/index2.html

Liu L, Oza S, Hogan D, et al. Global, regional, and national causes of under-5 mortality in 2000-15: an updated systematic analysis with implications for the Sustainable Development Goals. Lancet 2016.
https://www.ncbi.nlm.nih.gov/pubmed/27839855

*Household status*

Poverty/food security

| | **2020** | **2021** | **2022** | **2023** | **2024** | **2025** | **2026** | **2027** | **2028** | **2029** | **2030** |
|---|---|---|---|---|---|---|---|---|---|---|---|
| **Percentage of the population that is food insecure** | 22.6% | 22.6% | 22.6% | 22.6% | 22.6% | 22.6% | 22.6% | 22.6% | 22.6% | 22.6% | 22.6% |

Source: SIVESNU 2015

#### Intervention Coverages

*Notes*

- Abbreviations
    - HT = Historical trends
    - GC = Great Crusade
- Coverage levels in the Great Crusade model remain static after 2024.
- The Great Crusade fertility and aspirational model are not included in the tables for clarity.
    - The Great Crusade fertility model has the same coverages as the Great Crusade model.
    - Coverage levels in the Aspirational model reach 90.0% in 2024 and remain static till 2030.
- Coverages are linearly interpolated.

*Pregnancy*

Nutritional

| Input | Baseline (2020) | HT (2024) | HT (2030) | GC (2024) | Source |
|---|---|---|---|---|---|
| **Calcium supplementation in pregnancy** | No data | | | | |
| **Multiple micronutrient supplementation in pregnancy** | 82.3% | 82.3% | 82.3% | 90.0% | SIVESNU 2018 |

*Breastfeeding*

Breastfeeding Prevalence

| Input | Baseline (2020) | HT (2024) | HT (2030) | GC (2024) | Source |
|---|---|---|---|---|---|
| **Prevalence of early initiation of breastfeeding** | 66.2% | 68.6% | 72.0% | 71.2% | Projected value calculated by authors using historical ENSMI data. |

| | <1 month (2020) | HT (2024) | HT (2030) | GC (2024) |
|---|---|---|---|---|
| **Exclusive** | 63.7% | 65.1% | 68.4% | 67.7% |
| **Predominant** | 13.5% | 12.9% | 11.5% | 12.3% |
| **Partial** | 19.1% | 18.5% | 17.1% | 17.4% |
| **None** | 3.7% | 3.5% | 3.0% | 2.6% |
| **Total** | 100.0% | 100.0% | 100.0% | 100.0% |

| | 1-5 months (2020) | HT (2024) | HT (2030) | GC 2024) |
|---|---|---|---|---|
| **Exclusive** | 52% | 53.9% | 56.7% | 56.0% |
| **Predominant** | 16.9% | 14.7% | 14.7% | 15.0% |
| **Partial** | 25.8% | 23.5% | 23.5% | 23.9% |
| **None** | 5.3% | 5.1% | 5.1% | 5.1% |
| **Total** | 100.0% | 100.0% | 100.0% | 100.0% |

| | 6-11 months (2020) | HT (2024) | HT (2030) | GC (2024) |
|---|---|---|---|---|
| **Any type** | 90.0% | 90.0% | 90.0% | 90.0% |
| **None** | 10.0% | 10.0% | 10.0% | 10.0% |
| **Total** | 100.0% | 100.0% | 100.0% | 100.0% |

| | 12-23 months (2020) | HT (2024) | HT (2030) | GC (2024) |
|---|---|---|---|---|
| **Any type** | 71.3% | 71.6% | 72.1% | 71.6% |
| **None** | 28.7% | 28.4% | 27.9% | 28.4% |
| **Total** | 100.0% | 100.0% | 100.0% | 100.0% |

Source: Projected value calculated by authors using historical ENSMI data.

*Preventative*

Food and supplementation

| Input | Baseline (2020) | HT (2024) | HT (2030) | GC (2024) | Source |
|---|---|---|---|---|---|
| **Comp. feeding (only education)** | 62.6% | 62.6% | 62.6% | 72.6% | ENSMI 2014-2015 |
| **Comp. feeding (feeding & education)** | 62.6% | 62.6% | 62.% | 72.6% | ENSMI 2014-2015 |
| **Vitamin A supplementation for children** | 26.0% | 26.0% | 26.0% | 51.0% | Vitamin A Deficiency in Children - UNICEF Data.https://data.unicef.org/topic/nutrition/vitamin-a-deficiency/. Published 2019 |
| **Zinc supplementation for children** | 86.1% | 86.1% | 86.1% | 90.0% | SIVESNU 2018 |

WASH (water, sanitation and hygiene)

| Input | Baseline (2020) | HT (2024) | HT (2030) | GC (2024) | Source |
|---|---|---|---|---|---|
| **Basic sanitation** | 65.4% | 65.7% | 66.2% | 90.0% | Projected value calculated by authors using historical data from: WHO/UNICEF Joint Monitoring Program (JMP) for Water Supply and Sanitation (https://washdata.org/). Data are available for all countries from 1996 to 2015. |
| **Point-of-use water filter or piped in water** | 89.2% | 91.4% | 93.9% | 91.4% | |
| **Handwashing with soap** | 76.9% | 77.1% | 77.4% | 90.0% | |

*Vaccines*

| Input | Baseline (2020) | HT (2024) | HT (2030) | GC (2024) | Source |
|---|---|---|---|---|---|
| **Rotavirus (2 doses)** | 88.4% | 90.0% | 90.0% | 90.0% | Projected value calculated by authors using historical data from ENSMI. |

*Curative*

| Input | Baseline (2020) | Source |
|---|---|---|
| **Kangaroo mother care (KMC)** | No data | |

*Fertility Risks*

#### Maternal age

| **Less than 18 years old** | **2020*** | **2030*** | **GC (fertility) 2030** |
|---|---|---|---|
| **First birth** | 8.0% | 7.8% | 0.0% |
| **Second and third birth** | 0.9% | 0.6% | 0.0% |
| **Greater than third birth** | 0.0% | 0.0% | 0.0% |
| **18 to 34 years old** | | | |
| **First birth** | 26.5% | 32.1% | 39.9% |
| **Second and third birth** | 39.5% | 39.2% | 39.8% |
| **Greater than third birth** | 14.5% | 9.1% | 9.1% |
| **35 to 49 years old** | | | |
| **First birth** | 0.5% | 0.9% | 0.9% |
| **Second and third birth** | 3.3% | 5.9% | 5.9% |
| **Greater than third birth** | 6.8% | 4.4% | 4.4% |
| **Total** | 100% | 100% | 100.0% |

Source: Calculated by authors using microdata from ENSMI 2014-2015.

#### Birth intervals

| **Birth intervals** | **2020*** | **2030*** | **GC (fertility) 2030** |
|---|---|---|---|
| **First birth** | 35.4% | 43.3% | 43.3% |
| **Less than 18 months** | 3.0% | 2.0% | 0.0% |
| **18-23 months** | 6.6% | 4.3% | 0.0% |
| **24 months or more** | 55.0% | 50.4% | 56.7% |
| **Total** | 100.0% | 100% | 100% |

Source: Calculated by authors using microdata from ENSMI 2014-2015.

*Note: The historical trends, Great Crusade, and aspirational models have the same fertility risk.

### MODEL OUTPUTS

#### Stunting Prevalence

| Stunting Prevalence Median % (95% CI) | 2020 | 2021 | 2022 | 2023 | 2024 | 2025 | 2026 | 2027 | 2028 | 2029 | 2030 |
|---|---|---|---|---|---|---|---|---|---|---|---|
| **Historical Trends** | 46.48 (46.48 - 46.48) | 46.51 (46.49 - 46.52) | 46.51 (46.46 - 46.55) | 46.38 (46.3 - 46.43) | 46.39 (46.26 - 46.47) | 46.25 (46.08 - 46.36) | 46.29 (46.08 - 46.43) | 46.19 (45.94 - 46.36) | 46.11 (45.81 - 46.3) | 46.18 (45.84 - 46.4) | 46.09 (45.71 - 46.33) |
| **Great Crusade** | 46.48 (46.48 - 46.48) | 46.37 (46.3 - 46.42) | 46.16 (45.99 - 46.27) | 45.75 (45.46 - 45.93) | 45.41 (44.98 - 45.68) | 45.03 (44.5 - 45.37) | 44.91 (44.3 - 45.3) | 44.72 (44.06 - 45.15) | 44.62 (43.93 - 45.07) | 44.71 (43.99 - 45.17) | 44.63 (43.89 - 45.11) |
| **Great Crusade (fertility)** | 46.48 (46.48 - 46.48) | 46.37 (46.29 - 46.42) | 46.16 (45.96 - 46.28) | 45.74 (45.4 - 45.95) | 45.39 (44.88 - 45.71) | 45.01 (44.37 - 45.41) | 44.87 (44.13 - 45.34) | 44.68 (43.86 - 45.2) | 44.58 (43.69 - 45.13) | 44.66 (43.71 - 45.24) | 44.57 (43.57 - 45.18) |
| **Aspirational** | 46.48 (46.48 - 46.48) | 46.25 (46.12 - 46.33) | 45.81 (45.49 - 46.02) | 45.06 (44.51 - 45.44) | 44.28 (43.45 - 44.85) | 43.56 (42.52 - 44.28) | 43.19 (42.01 - 44.02) | 42.88 (41.61 - 43.78) | 42.75 (41.44 - 43.68) | 42.83 (41.5 - 43.77) | 42.76 (41.41 - 43.71) |

#### Stunting Cases Averted

| Stunting Cases Averted | 2020 | 2021 | 2022 | 2023 | 2024 | 2025 | 2026 | 2027 | 2028 | 2029 | 2030 |
|---|---|---|---|---|---|---|---|---|---|---|---|
| **Historical Trends** | 0 (0 - 0) | 328 (126 - 680) | 757 (228 - 1,590) | 1,381 (457 - 2,803) | 1,997 (646 - 4,196) | 2,640 (766 - 5,659) | 3,307 (924 - 7,087) | 3,912 (1,038 - 8,418) | 4,479 (1,148 - 9,662) | 5,010 (1,272 - 10,836) | 5,496 (1,362 - 11,934) |
| **Great Crusade** | 0 (0 - 0) | 2,938 (2,042 - 4,290) | 7,254 (5,128 - 10,384) | 12,878 (9,246 - 18,153) | 19,684 (14,292 - 27,303) | 24,308 (17,683 - 33,587) | 27,584 (20,082 - 38,095) | 29,551 (21,413 - 40,947) | 30,178 (2,1691 - 42,078) | 30,345 (21,663 - 42,546) | 30,313 (21,466 - 42,772) |
| **Great Crusade (fertility)** | 0 (0 - 0) | 2,973 (2,000 - 4,462) | 7,324 (5,022 - 10,767) | 13,027 (9,041 - 18,820) | 19,884 (13,987 - 28,356) | 24,637 (17,254 - 35,116) | 28,038 (19,538 - 40,140) | 30,098 (20,724 - 43,549) | 30,853 (20,838 - 45,302) | 31,120 (20,648 - 46,349) | 31,213 (20,277 - 47,222) |
| **Aspirational** | 0 (0 - 0) | 5,254 (3,572 - 7,631) | 13,760 (9,598 - 19,489) | 25,324 (17,975 - 35,240) | 39,632 (28,567 - 54,259) | 49,992 (36,174 - 68,041) | 57,128 (41,473 - 77,443) | 61,107 (44,284 - 82,853) | 62,067 (44,754 - 84,412) | 62,069 (44,665 - 84,557) | 61,625 (44,172 - 84,193) |

#### Stunting Cases Averted by Intervention

##### *Historical Trends*

| Stunting Cases Averted | 2020 | 2021 | 2022 | 2023 | 2024 | 2025 | 2026 | 2027 | 2028 | 2029 | 2030 |
|---|---|---|---|---|---|---|---|---|---|---|---|
| **Birth intervals** | 0 (0 - 0) | 0 (0 - 62) | 9 (0 - 161) | 17 (0 - 281) | 30 (0 - 466) | 44 (0 - 761) | 54 (0 - 939) | 66 (0 - 1,140) | 80 (0 - 1,261) | 91 (0 - 1,432) | 101 (0 - 1,609) |
| **Breastfeeding** | 0 (0 - 0) | 16 (3 - 52) | 42 (2 - 128) | 95 (8 - 254) | 144 (10 - 386) | 217 (14 - 563) | 266 (16 - 696) | 332 (19 - 867) | 407 (26 - 1,059) | 440 (24 - 1,154) | 499 (27 - 1,304) |
| **Filtered water/piped in water** | 0 (0 - 0) | 312 (155 - 563) | 677 (275 - 1,240) | 1,218 (500 - 2,167) | 1,728 (703 - 3,157) | 2,253 (858 - 4,091) | 2,812 (1,024 - 5,112) | 3,310 (1,164 - 6,012) | 3,761 (1,284 - 6,886) | 4,200 (1,423 - 7,697) | 4,594 (1,547 - 8,422) |
| **Basic sanitation** | 0 (0 - 0) | 0 (0 - 0) | 15 (5 - 28) | 35 (13 - 65) | 58 (21 - 109) | 86 (29 - 160) | 113 (36 - 210) | 141 (44 - 262) | 144 (44 - 269) | 170 (51 - 319) | 194 (58 - 365) |
| **Hand washing with soap** | 0 (0 - 0) | 0 (0 - 0) | 14 (5 - 29) | 16 (6 - 33) | 36 (13 - 74) | 40 (14 - 82) | 63 (21 - 128) | 65 (21 - 134) | 89 (27 - 183) | 113 (34 - 232) | 113 (34 - 232) |
| **Rotavirus vaccine** | 0 (0 - 0) | 3 (2 - 3) | 3 (2 - 3) | 3 (2 - 3) | 2 (2 - 3) | 2 (2 - 3) | 2 (1 - 3) | 2 (1 - 3) | 2 (1 - 3) | 2 (1 - 3) | 2 (1 - 3) |

*Great Crusade*

| Stunting Cases Averted | 2020 | 2021 | 2022 | 2023 | 2024 | 2025 | 2026 | 2027 | 2028 | 2029 | 2030 |
|---|---|---|---|---|---|---|---|---|---|---|---|
| **Micronutrient supplementation (iron and multiple micronutrients)** | 0 (0 - 0) | 17 (8 - 30) | 48 (26 - 83) | 89 (49 - 152) | 138 (78 - 232) | 178 (102 - 293) | 194 (111 - 319) | 203 (116 - 336) | 204 (116 - 340) | 200 (113 - 333) | 197 (109 - 329) |
| **Birth intervals** | 0 (0 - 0) | 6 (0 - 69) | 14 (0 - 176) | 23 (0 - 306) | 39 (0 - 496) | 63 (0 - 901) | 80 (0 - 1,143) | 96 (0 - 1,375) | 113 (0 - 1,471) | 124 (0 - 1,624) | 134 (0 - 1,786) |
| **Breastfeeding** | 0 (0 - 0) | 45 (4 - 104) | 121 (7 - 279) | 223 (14 - 505) | 363 (36 - 789) | 467 (45 - 997) | 512 (51 - 1,089) | 538 (54 - 1,144) | 540 (54 - 1,155) | 530 (53 - 1,132) | 520 (51 - 1,114) |
| **Complementary feeding** | 0 (0 - 0) | 758 (737 - 766) | 2,199 (2,147 - 2,222) | 4,340 (4,241 - 4,389) | 7,172 (6,997 - 7,266) | 9,214 (9,011 - 9,313) | 10,640 (10,380 - 10,757) | 11,389 (11,061 - 11,527) | 11,532 (11,113 - 11,698) | 11,455 (10,981 - 11,625) | 11,265 (10,745 - 11,441) |
| **Zinc supplementation** | 0 (0 - 0) | 438 (411 - 478) | 924 (865 - 1,004) | 1,482 (1,376 - 1,602) | 2,105 (1,948 - 2,278) | 2,257 (2,084 - 2,451) | 2,423 (2,228 - 2,632) | 2,509 (2,298 - 2,728) | 2,503 (2,277 - 2,725) | 2,488 (2,253 - 2,714) | 2,454 (2,213 - 2,677) |
| **Filtered water/piped in water** | 0 (0 - 0) | 350 (201 - 567) | 764 (445 - 1,230) | 1,334 (781 - 2,132) | 1,905 (1,121 - 3,028) | 2,668 (1,569 - 4,204) | 3,452 (2,030 - 5,436) | 4,069 (2,385 - 6,405) | 4,536 (2,644 - 7,166) | 4,938 (2,866 - 7,799) | 5,308 (3,064 - 8,387) |
| **Basic sanitation** | 0 (0 - 0) | 885 (449 - 1,467) | 2,116 (1,093 - 3,476) | 3,590 (1,870 - 5,861) | 5,291 (2,773 - 8,598) | 6,281 (3,300 - 10,102) | 6,822 (3,584 - 10,960) | 7,124 (3,730 - 11,438) | 7,128 (3,712 - 11,485) | 7,036 (3,648 - 11,340) | 6,920 (3,567 - 11,156) |
| **Hand washing with soap** | 0 (0 - 0) | 437 (229 - 805) | 1,044 (551 - 1,898) | 1,772 (937 - 3,193) | 2,611 (1,385 - 4,676) | 3,097 (1,639 - 5,484) | 3,364 (1,778 - 5,952) | 3,513 (1,850 - 6,212) | 3,515 (1,840 - 6,237) | 3,470 (1,809 - 6,159) | 3,412 (1,769 - 6,059) |
| **Rotavirus vaccine** | 0 (0 - 0) | 3 (2 - 3) | 3 (2 - 3) | 2 (2 - 3) | 2 (2 - 3) | 2 (2 - 3) | 2 (2 - 3) | 2 (2 - 3) | 2 (2 - 3) | 2 (2 - 2) | 2 (2 - 2) |

*Great Crusade (fertility)*

| Stunting Cases Averted | 2020 | 2021 | 2022 | 2023 | 2024 | 2025 | 2026 | 2027 | 2028 | 2029 | 2030 |
|---|---|---|---|---|---|---|---|---|---|---|---|
| **Micronutrient supplementation (iron and multiple micronutrients)** | 0 (0 - 0) | 9 (3 - 15) | 25 (11 - 42) | 46 (22 - 76) | 71 (36 - 116) | 91 (47 - 145) | 99 (51 - 157) | 104 (53 - 165) | 105 (52 - 166) | 103 (51 - 163) | 101 (49 - 161) |
| **Maternal age and birth order** | 0 (0 - 0) | 14 (8 - 15) | 41 (28 - 42) | 75 (56 - 76) | 118 (92 - 119) | 183 (150 - 186) | 247 (204 - 254) | 300 (244 - 309) | 344 (273 - 356) | 375 (296 - 388) | 409 (319 - 425) |
| **Birth intervals** | 0 (0 - 0) | 17 (0 - 235) | 48 (0 - 650) | 88 (0 - 1191) | 138 (0 - 1835) | 224 (0 - 2906) | 298 (0 - 3821) | 369 (0 - 4722) | 431 (0 - 5512) | 482 (0 - 6173) | 536 (0 - 6867) |
| **Breastfeeding** | 0 (0 - 0) | 46 (4 - 105) | 123 (7 - 277) | 226 (14 - 500) | 366 (34 - 778) | 470 (43 - 974) | 516 (48 - 1,060) | 542 (50 - 1,110) | 546 (49 - 1,118) | 536 (48 - 1,099) | 527 (47 - 1,084) |
| **Complementary feeding** | 0 (0 - 0) | 761 (725 - 767) | 2,196 (2,096 - 2,220) | 4,337 (4,134 - 4,380) | 7,169 (6,820 - 7,251) | 9,170 (8,739 - 9,291) | 10,571 (10,013 - 10,719) | 11,302 (10,620 - 11,477) | 11,441 (10,609 - 11,615) | 11,377 (10,411 - 11,558) | 11,211 (10,108 - 11,379) |
| **Zinc supplementation** | 0 (0 - 0) | 438 (411 - 478) | 924 (863 - 1,001) | 1,482 (1,364 - 1,598) | 2,107 (1,919 - 2,281) | 2,260 (2,042 - 2,453) | 2,425 (2,176 - 2,627) | 2,514 (2,237 - 2,716) | 2,510 (2,208 - 2,712) | 2,498 (2,172 - 2,699) | 2,466 (2,121 - 2,666) |
| **Filtered water/piped in water** | 0 (0 - 0) | 352 (195 - 568) | 767 (431 - 1,221) | 1,340 (754 - 2,107) | 1,911 (1,083 - 2,985) | 2,678 (1,510 - 4,120) | 3,467 (1,946 - 5,305) | 4,084 (2,268 - 6,235) | 4,558 (2,490 - 6,961) | 4,965 (2,685 - 7,588) | 5,345 (2,853 - 8,178) |
| **Basic sanitation** | 0 (0 - 0) | 891 (433 - 1,470) | 2,124 (1,055 - 3,450) | 3,606 (1,801 - 5,790) | 5,309 (2,674 - 8,470) | 6,301 (3,170 - 9,901) | 6,846 (3,429 - 10,708) | 7,144 (3,541 - 11,151) | 7,157 (3,488 - 11,174) | 7,071 (3,411 - 11,050) | 6,964 (3,315 - 10,894) |
| **Hand washing with soap** | 0 (0 - 0) | 440 (221 - 806) | 1,049 (533 - 1,884) | 1,780 (905 - 3,155) | 2,619 (1,338 - 4,608) | 3,109 (1,577 - 5,373) | 3,378 (1,703 - 5,808) | 3,525 (1,758 - 6,046) | 3,531 (1,732 - 6,058) | 3,489 (1,694 - 5,991) | 3,436 (1,647 - 5,906) |
| **Rotavirus vaccine** | 0 (0 - 0) | 3 (2 - 3) | 3 (2 - 3) | 2 (2 - 3) | 2 (2 - 3) | 2 (2 - 3) | 2 (2 - 3) | 2 (2 - 3) | 2 (2 - 2) | 2 (2 - 2) | 2 (2 - 2) |

*Aspirational*

| Stunting Cases Averted | 2020 | 2021 | 2022 | 2023 | 2024 | 2025 | 2026 | 2027 | 2028 | 2029 | 2030 |
|---|---|---|---|---|---|---|---|---|---|---|---|
| **Micronutrient supplementation (iron and multiple micronutrients)** | 0 (0 - 0) | 11 (0 - 20) | 30 (12 - 53) | 55 (24 - 96) | 83 (39 - 145) | 106 (53 - 183) | 115 (58 - 198) | 121 (61 - 207) | 121 (61 - 209) | 119 (60 - 205) | 117 (58 - 202) |
| **Birth intervals** | 0 (0 - 0) | 7 (0 - 90) | 17 (0 - 222) | 28 (0 - 381) | 46 (0 - 608) | 74 (0 - 1,096) | 94 (0 - 1,384) | 112 (0 - 1,660) | 132 (0 - 1,771) | 145 (0 - 1,953) | 158 (0 - 2,148) |
| **Breastfeeding** | 0 (0 - 0) | 638 (121 - 1,416) | 1,650 (281 - 3,602) | 3,008 (526 - 6,425) | 4,642 (853 - 9,695) | 5,820 (1,008 - 12,077) | 6,550 (1,193 - 13,464) | 6,936 (1,281 - 14,201) | 6,979 (1,291 - 14,305) | 6,887 (1,287 - 14,074) | 6,766 (1,263 - 13,813) |
| **Complementary feeding** | 0 (0 - 0) | 2,337 (2,151 - 2,479) | 6,739 (6,247 - 7,074) | 13,239 (12,278 - 13,912) | 21,819 (20,181 - 22,987) | 28,230 (26,136 - 29,564) | 32,666 (30,148 - 34,167) | 34,976 (32,187 - 36,520) | 35,358 (32,418 - 37,022) | 35,154 (32,187 - 36,812) | 34,611 (31,645 - 36,224) |
| **Zinc supplementation** | 0 (0 - 0) | 451 (418 - 499) | 989 (905 - 1,099) | 1,612 (1,444 - 1,809) | 2,304 (2,044 - 2,601) | 2,492 (2,191 - 2,826) | 2,682 (2,346 - 3,044) | 2,782 (2,426 - 3,158) | 2,773 (2,410 - 3,150) | 2,760 (2,395 - 3,135) | 2,725 (2,363 - 3,094) |
| **Filtered water/piped in water** | 0 (0 - 0) | 378 (202 - 626) | 841 (458 - 1,382) | 1,470 (808 - 2,410) | 2,081 (1,158 - 3,388) | 2,928 (1,639 - 4,737) | 3,789 (2,127 - 6,121) | 4,463 (2,502 - 7,199) | 4,972 (2,776 - 8,031) | 5,415 (3,022 - 8,731) | 5,829 (3,242 - 9,386) |
| **Basic sanitation** | 0 (0 - 0) | 951 (447 - 1,605) | 2,329 (1,122 - 3,906) | 3,949 (1,926 - 6,605) | 5,782 (2,861 - 9,607) | 6,895 (3,445 - 11,382) | 7,488 (3,754 - 12,351) | 7,813 (3,912 - 12,873) | 7,814 (3,897 - 12,889) | 7,717 (3,845 - 12,711) | 7,600 (3,774 - 12,501) |
| **Hand washing with soap** | 0 (0 - 0) | 477 (231 - 893) | 1,158 (571 - 2,147) | 1,949 (967 - 3,598) | 2,852 (1,430 - 5,225) | 3,399 (1,711 - 6,174) | 3,691 (1,862 - 6,696) | 3,852 (1,940 - 6,977) | 3,852 (1,932 - 6,984) | 3,805 (1,906 - 6,888) | 3,747 (1,872 - 6,775) |
| **Rotavirus vaccine** | 0 (0 - 0) | 3 (2 - 3) | 3 (2 - 3) | 3 (2 - 3) | 3 (2 - 3) | 3 (2 - 3) | 3 (2 - 3) | 3 (2 - 3) | 2 (2 - 3) | 2 (2 - 3) | 2 (2 - 3) |
